# Predicting prenatal depression and assessing model bias using machine learning models

**DOI:** 10.1101/2023.07.17.23292587

**Authors:** Yongchao Huang, Suzanne Alvernaz, Sage J. Kim, Pauline Maki, Yang Dai, Beatriz Peñalver Bernabé

## Abstract

Perinatal depression (PND) is one of the most common medical complications during pregnancy and postpartum period, affecting 10-20% of pregnant individuals. Black and Latina women have higher rates of PND, yet they are less likely to be diagnosed and receive treatment. Machine learning (ML) models based on Electronic Medical Records (EMRs) have been effective in predicting postpartum depression in middle-class White women but have rarely included sufficient proportions of racial and ethnic minorities, which contributed to biases in ML models for minority women. Our goal is to determine whether ML models could serve to predict depression in early pregnancy in racial/ethnic minority women by leveraging EMR data. We extracted EMRs from a hospital in a large urban city that mostly served low-income Black and Hispanic women (N=5,875) in the U.S. Depressive symptom severity was assessed from a self-reported questionnaire, PHQ-9. We investigated multiple ML classifiers, used Shapley Additive Explanations (SHAP) for model interpretation, and determined model prediction bias with two metrics, Disparate Impact, and Equal Opportunity Difference. While ML model (Elastic Net) performance was low (ROCAUC=0.67), we identified well-known factors associated with PND, such as unplanned pregnancy and being single, as well as underexplored factors, such as self-report pain levels, lower levels of prenatal vitamin supplement intake, asthma, carrying a male fetus, and lower platelet levels blood. Our findings showed that despite being based on a sample mostly composed of 75% low-income minority women (54% Black and 27% Latina), the model performance was lower for these communities. In conclusion, ML models based on EMRs could moderately predict depression in early pregnancy, but their performance is biased against low-income minority women.

## INTRODUCTION

Perinatal depression (PND), depression during pregnancy and up to one year postpartum, is one of the most common complications during the perinatal period (1). In the United States (U.S.), the rate of PND is 10-20% (1) and has increased more than 3-fold from 2000 to 2015 (2, 3). The rates of PND among Black women and Latinas are 2-to 5-fold higher than non-Hispanic White Women (4–8). In our own longitudinal studies in low-income women of color, the rate of PND is 23%, while the U.S. average rate is 12%. During the COVID-19 pandemic, the prevalence of PND has risen to 27%-32% (9–11), highlighting the importance of environmental stressors in these mood disorders (12). PND confers significant obstetric risks of low birth weight (13), preterm labor (13, 14), higher maternal morbidity and mortality, longer hospital stays post-delivery and higher delivery costs (15), lower initiation and duration of breastfeeding (16), and poor maternal-fetal attachment (17). Infants born from women with PND have increased risks of stunted growth, inadequate cognitive development, altered stress response, underdeveloped social-emotional behavior, and future mental disorders (15, 18–23). In extreme cases, PND can lead to suicide, a leading cause of maternal mortality in the first year after delivery (24).

Multiple individual-level factors have been linked to increased PND risk, such as lack of a partner and social support (25), unplanned pregnancy (26), young age (27), prior history of depression or trauma (27) and adverse childhood experiences (28, 29). Despite a higher prevalence of depression during pregnancy in Black women and Latinas, they are less likely to be screened (30–32) and to share their PND symptoms (33–35). Providers commonly screen for depression at least once during pregnancy and postpartum (36). Common PND screening methods include self-reported questionnaires, such as Edinburgh Postpartum Depression Score (EPDS) (37) and the Patient Health Questionnaire-9 (PHQ-9) (38). However, the utility of these self-reported questionnaires depends on the accurate disclosure of symptoms (33–35). For instance, Black women might share their PND symptoms with family but are more reluctant to seek help compared to non-Hispanic White women due to social stigma (33, 39) and they might be concerned that they are viewed as lacking strength and resilience (40–42). Fear of the consequences of symptoms disclosure, such as losing the infant’s custody (33), and medical mistrust are also important factors for which pregnant individuals might decide to underreport or completely deny their symptoms (43). Furthermore, how depressive symptoms are communicated might be influenced by cultural factors that EPDS or PHQ-9 might not be fully captured (43–47). For example, studies have shown that low-income urban Black women endorse lower levels of depressive symptoms severity in self-reported questionnaires than other populations (45–47), despite having the same clinical diagnosis, hence leading to more false negative results in Black women compared with other groups.

Computational approaches, such as machine learning (ML) using Electronic Medical Records (EMRs), have been able to predict pregnancy outcomes, including gestational diabetes (48–50), preterm birth (51), and suicidal thoughts (52). EMR-based ML models for PND have generally focused on predicting postpartum depression (53–63), and rarely include racial and ethnic minorities (54, 59). In middle-class White women, EMR-based ML models to predict postpartum depression can perform relatively well, with Receiver Operating Characteristic Area Under the Curve (ROCAUC)>0.75, using various approaches such as random forest (RF) (54, 58, 59, 63–65), artificial neural networks (ANNs) (55, 56, 66), and logistic regression (54, 62, 67). However, the performance of EMR-based ML models for predicting depression in pregnancy for low-income minority women remains unexplored.

Biases in prediction performance exist in ML models, which refer to the disparate levels of model prediction of the outcome of interest for certain socio-demographic variables, called protected variables, such as sex, gender, age, race, ethnicity, or social-economic status (68). Since EMRs are not collected from well-designed balanced studies, the model performance can better predict or be biased towards groups more represented in EMRs. Also, some groups might have more available data (e.g., more clinical encounters, diagnostic tests), or their EMR information is of higher quality (69). White women are commonly disproportionally overrepresented in EMR, and consequently, ML models based on EMR have lower performance at predicting PND risk in Black and Latina women compared with White individuals (59). Other reasons for poor performance include the lower quality and quantity of EMRs for racial/ethnic minorities than those of White women because of lower access to care (59), e.g., no prior documented medical history in the EMRs; underutilization of health care due to work conditions; lack of childcare or transportation; reluctance to reveal certain sensitive information (33); and/or implicit bias by health care providers (33, 43). Thus, understanding the sources of bias in ML models is essential for the equitable prediction of PND risk in diverse populations.

Despite the negative consequences of depression during pregnancy for both the mother and the infant that disproportionally impact low-income women of color, no study has thus far used EMR information enriched in women of color from under-resourced communities to predict depression early in pregnancy and examined the model bias. Here, we constructed ML models to predict depression symptom severity early in pregnancy for an urban low-income women of color that received care in the same outpatient obstetric clinic, and we subsequently assessed model performance and racial and ethnic biases.

## MATERIALS AND METHODS

### Study population

EMRs were extracted from patients who received obstetric care at the University of Illinois in Chicago (UIC) Hospital Health Sciences System, UIC Medical Center (UIHealth), an urban academic hospital in Chicago, U.S. The population served at UIHealth is 51% non-Hispanic Black, 29% Hispanic, 9% non-Hispanic White, and 10% Asian and Native American combined. We extracted EMR from patients who were 18 years or older and received their obstetric care and delivered within UIHealth between 2014 to 2020. We obtained information on the patient’s socio-demographic characteristics, prior medical history, mental health assessments, health behaviors (alcohol use, smoking status, substance abuse, etc.), vital records, laboratory tests, medications, obstetric complications for the prior and current pregnancy, and delivery and fetus information (weight, length, sex, Apgar scores). Depressive symptom severity was assessed using a self-reported questionnaire PHQ-9 (38). The extracted data included 694 features and 5,875 patients. The entire data dictionary is available in **Table S1 and S2.** The EMR extraction was approved by the University of Illinois Institutional Review Board (IRB # 2020-0553).

### Preprocessing of EMR data

Patients that satisfy the following criteria were included in the downstream analysis: 1) patients with complete PHQ-9 (all 9 questions were recorded in the patient’s EMR); 2) patients that completed their first obstetric visit before 24 gestational weeks, as depressive symptoms later in pregnancy might be distinct from those experienced in the first and second trimesters; 3) patients with PHQ-9 scores between 1-4 or 9 and above. Patients with mild levels of depression (PHQ-9 scores between 5-8) or those who reported no depression symptoms (PHQ-9 score 0) were excluded to avoid incorporating possible false negatives in case the individual underreported or denied their symptoms, respectively. Patients with a PHQ-9 score higher than 9 were categorized as cases. Patients with a PHQ-9 score between 1-4 were categorized as controls.

Prescribed medications were grouped into 29 broad classes based on their most common use, mechanism of action, and targeted organ system. These included medications/prescriptions taken for diabetes, heart pathologies, anti-inflammatory/analgesics, cancer, gastrointestinal disorders, vitamins & supplements, thyroid disease, fertility, antibiotics, and mood/anxiety medications. The complete annotation table can be found **(Table S2)**. We omitted features that were missing in more than 60% of the included patients. This cutoff was selected based on the minimum number of samples that were required to adequately impute a feature without jeopardizing model performance and independently of the ML algorithm (**Figure S1**). All qualitative features (e.g., race, insurance, and medications) were hard-coded, and continuous features were transformed using min-max normalization. Missing data were imputed using *MICE* (version 3.15.0) (70). To robustly impute missing values, we generated a total of 50 imputed datasets, with 10 sampling iterations per imputed set. For continuous variables, missing data were estimated as the average across all the 50 imputed datasets, and for qualitative variables as the majority of a given category. To partially mitigate bias, ML models were trained by excluding self-reported race and ethnicity as well as results from genetic tests to assess ancestry. We also removed features that have non-causal associations with PND but may be implicitly related to the patient’s race and ethnicity, such as preferred language to communicate with their provider other than English.

### Machine learning model selection and training

We explored different ML models, including Random Forest, Elastic Net, and XGBoost, to identify the most suitable one for the prediction of cases and controls. Due to the imbalance between the numbers of cases (N = 657) and controls (N = 1,757), we selected the best model based on ROCAUC. The data were split into 80% for training and 20% for testing. The *GridSearchCV* function from the *sklearn* python package (version 1.2.0) was employed to determine the best hyperparameters for each model through a 5-fold cross-validated grid search using the training set.

For the most suitable method based on their ROCAUC, Elastic Net, we used the optimized hyperparameters to generate 20 models, each of which was trained using 400 controls and 400 cases randomly sampled from the training set. The accuracy, ROCAUC, positive predictive value (PPV), negative predictive value (NPV), and specificity were calculated for model evaluation. We also computed the harmonic mean of precision and recall (F1),

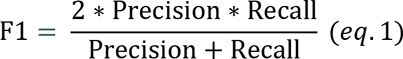

and the area under the Precision-Recall Curve (PRAUC),

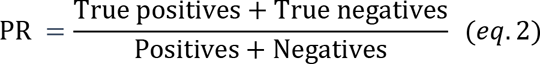

to further assess model performance. We fixed the sensitivity as 85% for each model to maximize the identification of cases—clinical objective.

### Identification of the most important features

To identify the importance and directionality with respect to depressive symptom severity of the EMR features, we calculated the Shapley values using SHAP (Shapley Additive Explanations) (71). The Shapley value, *ɸ*_*i*_, is defined as the estimated contribution of feature *i* in all samples to the depression outcome. For feature dependence analysis, we followed the procedure developed by Artzi and colleagues to convert the Shapley values into relative risk (RR) (48). Briefly, in the SHAP analysis, the log-odds of the predicted probability are calculated as *ɸ*_*i*_, then the predicted probability of a single feature *i* is

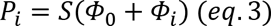

where,

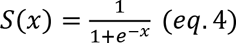

and *ɸ*_0_ is the ‘base’ Shapley value, i.e., the logit of the population prevalence (denoted as *P*_0_). Therefore,

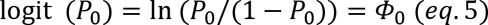

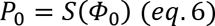

Here *P_0_*was set as 0.14, which was the prenatal depression prevalence estimated from the EMRs of the current study.

The relative risk (RR) of a single feature *i* was calculated as follows:

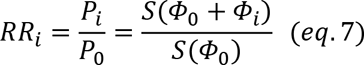

When calculating the *RR* of prenatal depression in relation to a set of features *A*, equation 7 can be extended as follows:

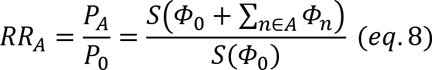

We employed the function *dependence_plot()* within the *shap* package (version 0.39.0) to determine the correlation between the most important features and the rest of the EMR variables based on their Shapley value.

### Statistical analysis

Differences between the groups were determined with the Mann-Whitney test and Bonferroni to correct for multiple comparisons. To establish the relationships between each feature and the outcome, we used Spearman correlation to estimate the direction of the effect.

### Bias assessments

We employed two common metrics to assess model bias, the disparate impact (DI) (72) and the equal opportunity difference (EOD) (73):

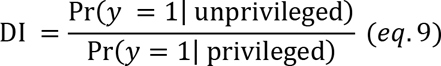

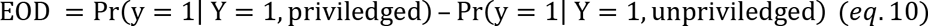

Here, *Y* and *y* denote the true and predicted outcome of depression (case=1, control=0), respectively. The privileged group was selected as the racial/ethnic group with the highest PRAUC and ROCAUC. In our study, the privileged group was non-Hispanic White women, and the unprivileged groups were non-Hispanic Black and Latina women.

## RESULTS

### EMRs were extracted from an understudied urban low-income woman of color

We extracted EMRs from a total of 5,875 patients who received obstetric care at an academic hospital affiliated with a public university in Chicago, U.S., from 2014-2020. After preprocessing the data (**Figure 1**, see methods for more detail), a total sample of 2,414 pregnant women was employed for downstream analysis (**Table 1**). Individuals were adults who received obstetric care and delivered in the public hospital and whose first obstetric visit for the given pregnancy was before 24 gestational weeks. Most of the available EMRs belongs to low-income women of color, 54% non-Hispanic Black and 27% non-Hispanic White women, and more than 72% of women were in federal aid health care plans (Medicaid and Medicare). We observed statistically significant differences between racial/ethnic groups **(Tables S3-S4**), with non-Hispanic Black women being more likely to be single (p-adj < 0.01), having an unplanned pregnancy (p-adj < 0.01), and being unemployed (p-adj < 0.01). We also detected statistically significant differences between cases and controls overall as well as when segregating the data by race/ethnicity, with women reporting high levels of depressive symptoms being more likely to have an unplanned pregnancy and to smoke independently of their race/ethnicity (p-adj < 0.01). The percentage of missing data for planned pregnancy, tobacco use, pain assessment scale was similar among races/ethnicity (**Figure S2c**).

**Figure 1.**
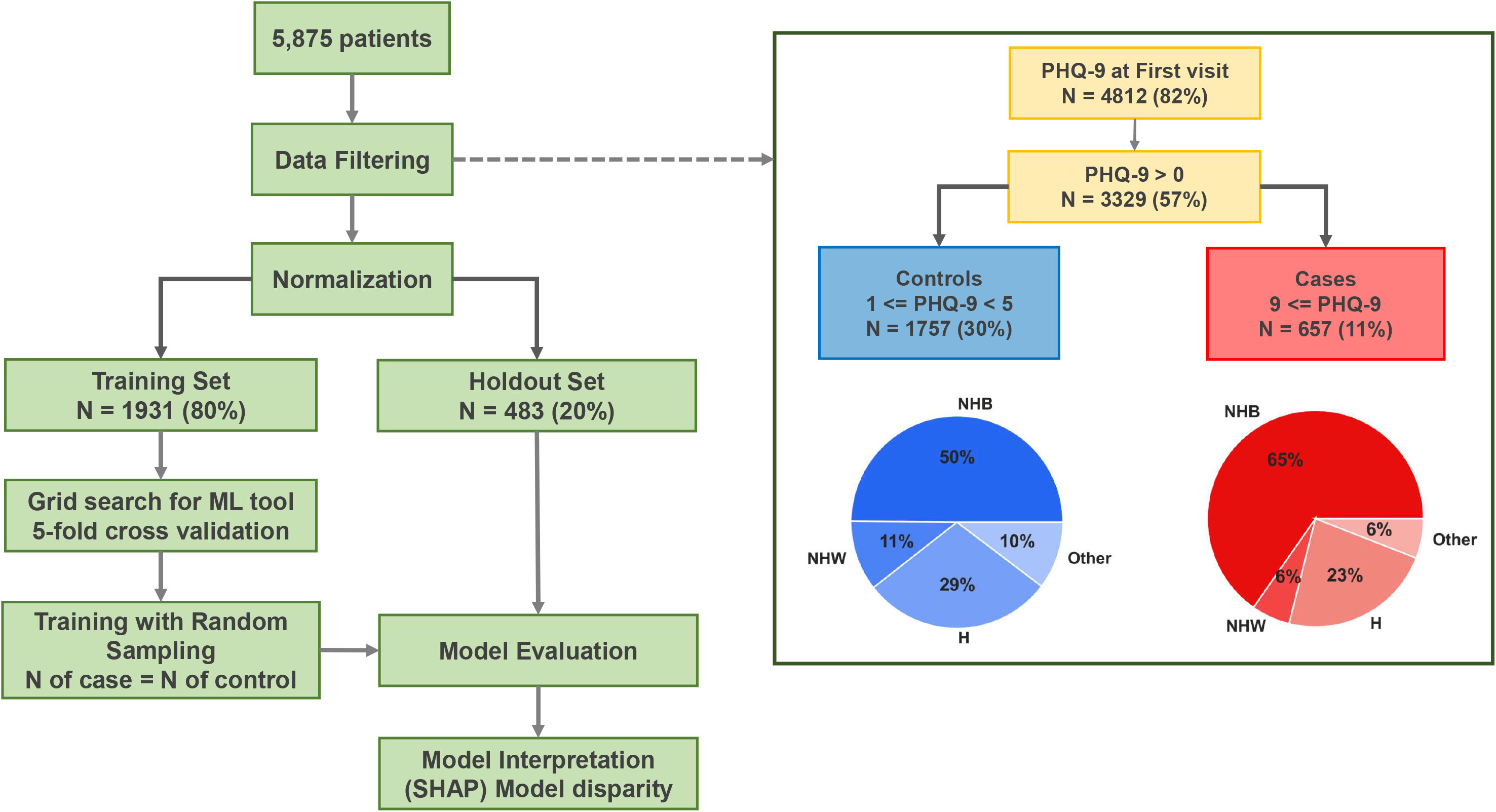
Overall workflow.

**Table 1.** Socio-demographic characteristics of the sample employed for model optimization.

| | | N, Percentage | | | | Mean ± s.d | | |
| --- | --- | --- | --- | --- | --- | --- | --- | --- |
|  | Race/Ethnicity | Single | Private Insurance | Planned pregnancy | Employed | Age | BMI | PHQ-9 |
| Cases<br>(N = 657) | All cases | 565 (86%) | 123 (19%) | 141 (22%) | 93 (14%) | 30 ± 6.4 | 30 ± 8.3 | 12 ± 3.5 |
| | NHB (429, 65%) | 398 (93%) | 64 (15%) | 57 (13%) | 47 (11%) | 29 ± 6.3 | 31 ± 9 | 12 ± 3.6 |
| | NHW (38, 6%) | 23 (61%) | 16 (42%) | 20 (53%) | 8 (21%) | 35 ± 6 | 27 ± 7 | 13 ± 4.0 |
| | H (151, 23%) | 121 (80%) | 26 (17%) | 46 (30%) | 19 (13%) | 30 ± 6.3 | 30 ± 7.6 | 12 ± 3.2 |
| | Other (39, 6%) | 23 (59%) | 17 (44%) | 18 (46%) | 4 (10%) | 32 ± 5.6 | 26 ± 5.3 | 13 ± 3.2 |
| Controls<br>(N = 1757) | All controls | 1258 (72%) | 548 (31%) | 717 (41%) | 405 (23%) | 32 ± 6.1 | 30 ± 8.3 | 2.4 ± 1.0 |
| | NHB (876; 50%) | 785 (90%) | 175 (20%) | 188 (21%) | 147 (17%) | 31 ± 5.9 | 31 ± 8.5 | 2.5 ± 1.0 |
| | NHW (188; 11%) | 71 (38%) | 124 (66%) | 143 (76%) | 56 (30%) | 35 ± 5.3 | 29 ± 7.3 | 2.4 ± 1.0 |
| | H (512, 29%) | 327 (64%) | 159 (31%) | 271 (53%) | 89 (17%) | 31 ± 6.3 | 30 ± 8.3 | 2.3 ± 1.0 |
| | Other (181; 10%) | 75 (41%) | 90 (50%) | 115 (64%) | 46 (25%) | 34 ± 6.1 | 27 ± 6.7 | 2.3 ± 1.0 |

Out of self-report PHQ-9 questions, questions pertaining to changes in appetite, sleep, and tiredness were commonly scored 1 or above in around 90% of the participants with the same proportions independently of race/ethnicity. In contrast, less than 5% of the individuals reported a score of 1 or above in feature related to suicide ideation and attempt, independently of their race or ethnicity (**Figure S3**).

### Elastic Net was the best model to predict depression in early pregnancy for our dataset

First, we determined the most adequate machine learning model to predict depressive symptom severity early in pregnancy in our sample. We explored multiple ML models, including Random Forest (74), XGBoost (75), and Elastic Net (76), and selected the model based on its performance at a fixed level of 85% sensitivity (**Figure 1)**. Both Elastic Net and Random Forest had similar performance when models were agnostic to patient’s race/ethnicity (**Table S5**). Given its lower computational time compared to the Random Forest, we performed all the downstream analyses with the Elastic Net models. There were no statistically significant differences between those Elastic Net models in which were agnostic to patient’s race and ethnicity (Model 1) and those that were not agnostic to patient’s racial and ethnical background (Model 2) (**Figure S4**).

### Identification of well-known and novel features associated with prenatal depressive symptom severity

To identify the features that were most predictive of depressive symptom severity early in pregnancy and the directionality of their associations, we determined their Shapley value. Based on game theory, Shapley value provides an estimate of the contribution made by each feature towards the overall prediction of the model. The top 20 most predictive features (based on their mean absolute Shapley values) included well-known socio-demographic factors associated with PND, such as having an unplanned pregnancy (77), being single (78), young (27), unemployed (79), on federal aid insurance (proxy for poverty) (80) and tobacco use (81) (**Figure 2a**). Yet, our model also identified features that have not been previously associated with depressive symptom severity in pregnancy, or just reported in a few studies. For instance, we discovered that elevated depressive symptoms were positively associated with self-reported levels of pain, an asthma diagnosis, carrying a male fetus (82), using antihistamines, analgesics, or antibiotics, and with lower platelet levels in blood (83, 84). Features related to preventive care, such as prenatal vitamin intake (85) and immunization against influenza and tetanus, were negatively correlated with depressive symptoms severity.

**Figure 2.**
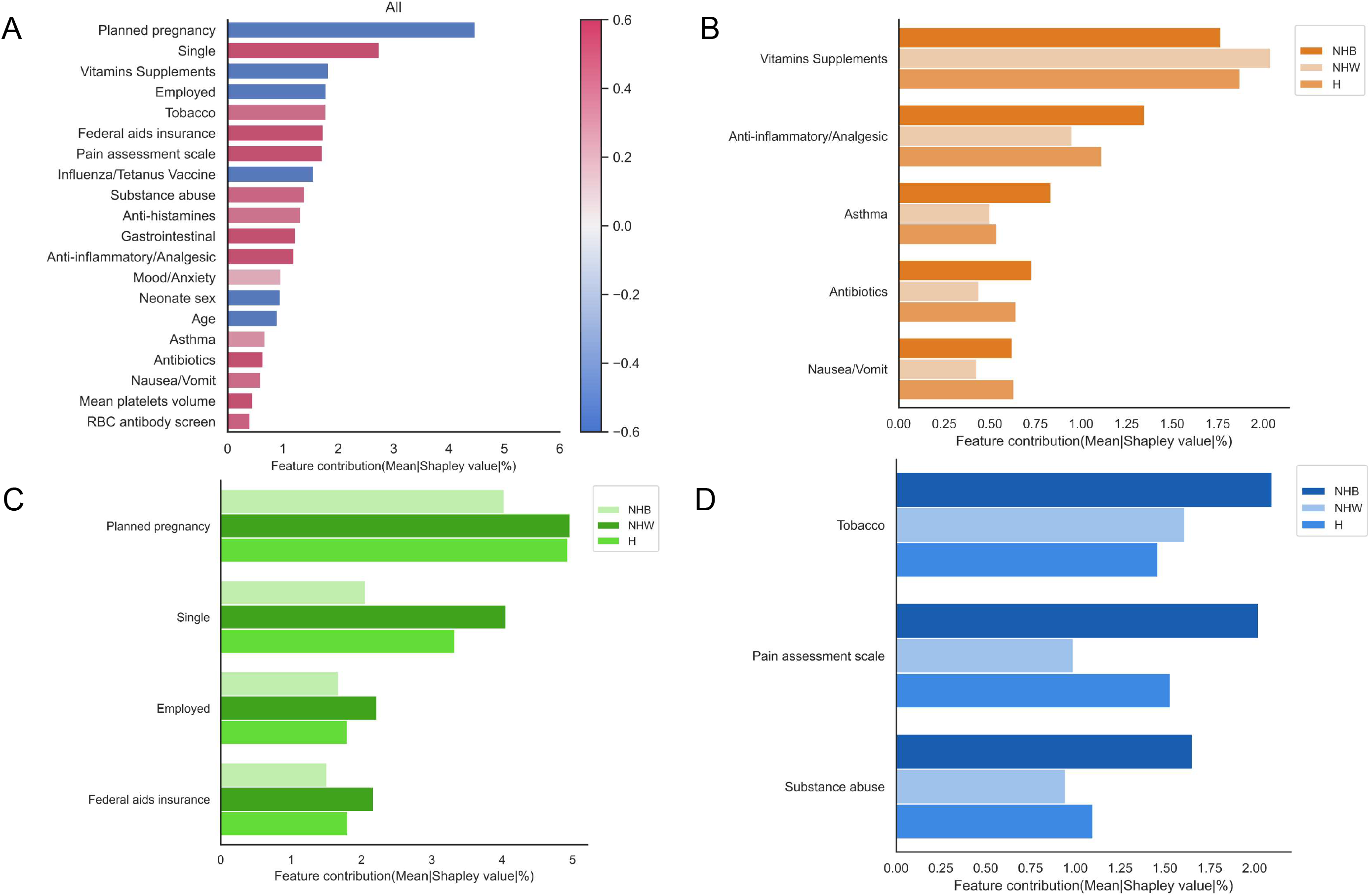
Multiple known and unexplored EMR features were associated with PND status. **a,** top 20 most important features based on their Shapley value to predict PND status. The color of the bar represents the correlation between each feature and PND status; red indicates a positive association with outcome and blue indicates a negative association. **b-d,** features whose importance is significantly different by race/ethnicity in terms of medication use **(b)**, socio-demographic factors **(c)**, substance use and pain assessment scale **(d)**. NHB: non-Hispanic Black. NHW: non-Hispanic White. H: Hispanic or Latina.

Inspection of the contribution of top significant features towards elevated levels of depressive symptoms as a function of patient’s race/ethnicity (**Figure 2b-d**), socio-demographic factors, such as being single, having an unplanned pregnancy, and low socio-economic status, were weaker predictors of depressive symptoms in non-Hispanic Black women compared to non-Hispanic White and Hispanic women (p < 0.01). However, tobacco use, infections, asthma diagnosis, and elevated self-report levels of pain had significantly higher contributions to the risk of prenatal depression in non-Hispanic Black women compared to the other two groups (p-value < 0.01). Compared to non-Hispanic White women, unemployment was a less predictive feature of prenatal depression risk in Black women, while experiencing nausea early in pregnancy had a significantly higher feature contribution (p-value < 0.05).

### Predicted relative risk of prenatal depression

Next, we determined the relative risk associated with most predicted features of depressive symptom severity based on their Shapley value (**Figure 3**). High self-reported pain levels were positively and linearly correlated with an increased risk of prenatal depression, independently of whether the patient was receiving any pain medication (**Figure 3a**). Of note, non-Hispanic Black individuals reported more often higher levels of pain (85%, score ≥ 6) than any other group (**Figure 3a)**. A history of medication for mood disorders is associated with an increased risk of perinatal depression (mean RR=1.18, sd=0.02, **Figure 3b**). As priorly reported (77, 78, 81), tobacco use (mean RR=1.09, sd=0.01, **Figure 3c**), unplanned pregnancy (RR=1.03, sd=0.01, **Figure 3d**), and being single (RR=1.02, sd=0.01, **Figure 3e**) are associated with increased the risk of perinatal depression.

**Figure 3.**
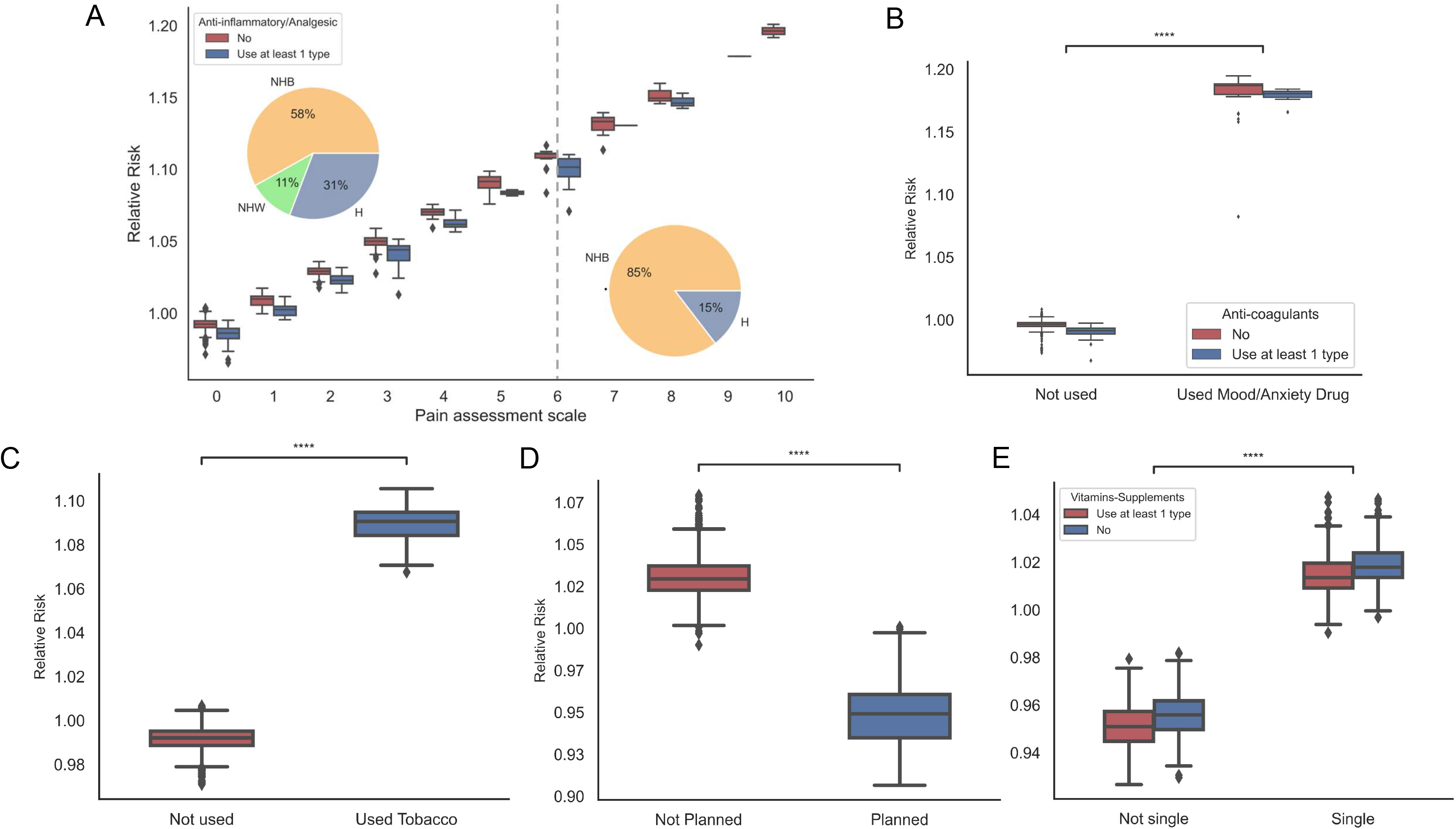
The predicted relative risk of top selected features. **a,** predicted relative risk of self-report pain level and its interaction with the use of anti-inflammatory/analgesic drugs. The pie chart represents the racial/ethnic distribution of pain assessment levels below and equal or above 6. **b-d,** predicted relative risk of planned pregnancy **(b)**, of being single and its interactions with vitamin supplement intake **(c),** of tobacco use **(d),** of mood/anxiety medication use and their interactions with anti-coagulants medication **(e)**.

### Despite the enrichment of minority low-income women of color, the model was biased against Black and Latina women

Finally, we examined whether ML models had the same predictive capability to determine PND status (cases versus controls) without using the individual’s race/ethnicity as part of the features for model training (**Figure 4a-b**). The model performance for any patient, independent of their racial/ethnic background, was moderate based on the Precision-Recall and Receiver Operating Characteristic Area under the Curve (PRAUC and ROCAUC, 50% and 66%, respectively). However, when stratifying the prediction by racial/ethnic groups, Elastic Net predictions of PND status for non-Hispanic White women were significantly higher than those for non-Hispanic Black women (PRAUC: 70%, 50%, and 40%; ROCAUC: 85%, 62%, and 70%, respectively; p<0.001). Further, the performance of the ML model for non-Hispanic White women was better than for non-Hispanic Black and Latina women when compared by specificity, accuracy, F1, PPV, and NPV (**Figure S4**).

**Figure 4.**
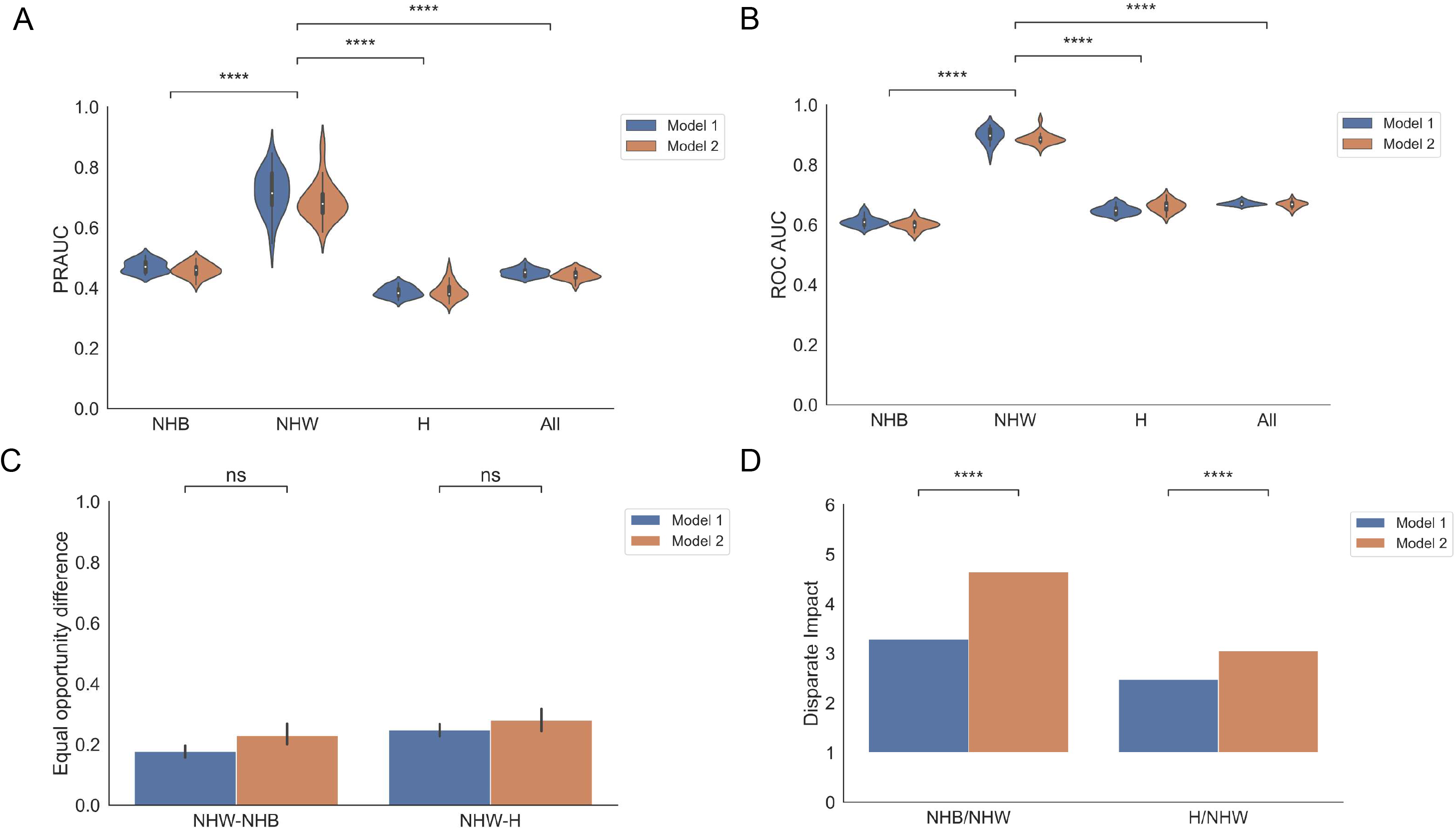
EMR-based ML models are biased against low-income minority women. **a,** area under the precision and recall curve for different racial/ethnic groups. **b,** the area under the receiver operating characteristic curve for different race/ethnicity. **c,** disparate impact values between unprivileged and privileged groups. Privileged group: non-Hispanic White women; unprivileged groups: non-Hispanic Black and Hispanic women. Model 1: trained without race/ethnicity label (blue). Model 2: trained with race/ethnicity label (orange).

To further estimate the inequality of model prediction or model bias, we calculated two common bias metrics: Equal Opportunity Difference (EOD), the true positive difference between the privileged and unprivileged group, and Disparity Impact (DI), the ratio of positive predictions between the privileged and unprivileged groups. Both DI and EOD values indicated that Elastic Net models had lower predictive performance in the unprivileged groups, non-Hispanic Black and Latina women, despite the model being trained with a sample mostly composed of low-income women of color. Removal of race/ethnicity as training features from the models moderately reduces model performance disparities as measured by DI, with more significant improvements for non-Hispanic Black women (p-value < 0.001, non-Hispanic Black versus no-Hispanic White women; p-value < 0.01, Hispanic versus no-Hispanic White women) (**Figure 4c-d**).

## DISCUSSION

In this study, we examined the capability of interpretable machine learning models to predict the risk of depression during early pregnancy in low-income women of color using electronic medical records. Our results demonstrated that EMR-based ML models were moderately predictive of depressive symptom severity in early pregnancy in low-income women of color. In addition, using the SHAP analysis, our results were interpretable in that we were able to identify factors that were associated with the risk of early prenatal depression including the directionality of their associations. Our models not only revealed well-known factors, such as unplanned pregnancy, prior history of medication use to treat mood disorders, or young age, but also captured novel or underexplored markers, such as self-report pain levels, asthma diagnosis, carrying a male fetus, or platelet levels in blood. Importantly, our results highlighted the significant performance disparity in model prediction between racial and ethnic groups, with women of color at greater disadvantages.

ML models to predict PND in racial and ethnic minorities are scarce. For instance, Zang and colleagues used a relatively well-balanced racial/ethnic group of urban pregnant individuals (18% non-Hispanic White, 27% Black) to validate their EMR-based ML models to predict postpartum depression (86). Park and colleagues focused on pregnant individuals at high-risk for mood disorders during pregnancy and postpartum that were covered by Medicaid (87). EMR-based ML models to predict postpartum depression have been developed in a wide variety of populations (e.g., Chinese (58), European (55, 56, 60, 63), non-Hispanic White women in the U.S.). These EMR-based ML models can perform relatively well (ROCAUC>0.75) using various ML approaches such as Random Forest(54, 63, 64, 86, 88), XGBoost (54, 57, 86), artificial neural networks (55, 56, 89), support-vector machines(56, 58, 64, 90) or logistic regression (54). However, previous studies have not demonstrated the predictive capability of EMR-based ML models to estimate PND status in low-income minority women. Our results filled this current gap and showed that EMR-based models trained with a sample primarily composed of low-income minority women moderately predict PND status. Yet, the Elastic Net model had a better prediction performance for the less prevalent group in the sample, non-Hispanic White women (ROCAUC > 0.85), compared with non-Hispanic Black women (ROCAUC > 0.6) and Latinas (ROCAUC > 0.65).

Our results are interpretable, a very important capability to identify markers that providers can intervene upon. Using SHAP analysis, we revealed several established markers that increased the risk of depression in early pregnancy, such as unplanned pregnancy (77), or being single (78). Our results also suggest that individuals that do not take vitamin supplements during early pregnancy may have an increased risk of depression. This observation aligns with results from a systematic review performed by Sparling and colleagues. The authors found that lower levels of folate, vitamin D, iron, selenium, zinc, fats, and fatty acids were associated with increased risks of perinatal depression (85), but those studies were limited in their population diversity, which either failed to include minority populations, or had limited sample sizes of minority groups.

Importantly, our analysis uncovered several new or underexplored markers that increased the predicted risk of depression during early pregnancy, such as self-report pain levels, mean platelet volume in blood (83, 84), carrying a male fetus (82), and having a diagnosis of asthma. Higher levels of pain have been linked to postpartum depression, and our findings suggest that this relationship also holds during the early stages of pregnancy(91) Similarly, patients with major depression exhibit a higher mean platelet volume (83, 84), but this phenomenon has not been investigated in the context of depression during pregnancy. Recently, Myers and colleagues reported that carrying a male fetus increases the odds of postnatal depression (82). While the mechanisms that link the fetus’s sex and depression status are unclear, women who carry male fetuses have lower estradiol levels in the blood than those carrying female fetuses (92) and lower levels of estrogens outside pregnancy are associated with higher levels of depressive symptoms (93). Also, women carrying male fetuses have higher levels of inflammatory markers, such as IL1B, in early pregnancy, and higher levels of inflammation have been linked to elevated levels of depression symptoms during pregnancy and postpartum (94, 95). Finally, our model indicates that an asthma diagnose increases the relative risk of early prenatal depression. While asthma may not be directly correlated with depression during pregnancy, air pollution is linked to an increased risk of asthma (96) and depression (97). Most of the women in our study live in underserved neighborhoods with high levels of outdoor pollutants and asthma prevalence. Thus, the associations between asthma diagnosis and depression during early pregnancy are most likely mediated by the negative impact of structural inequalities than by biological mechanisms.

Despite our original expectation, employing samples enriched in individuals from underserved populations is not enough to reduce EMR-based ML model performance bias. Our results revealed a significant disparity in performance among non-Hispanic White women, non-Hispanic Black, and Hispanic women, and it was not driven by the under-representation of the latter groups. Even though there was no significant difference in the overall performance between models trained with or without race and ethnicity, the non-Hispanic White women, the least represented group in the sample, were predicted significantly better than non-Hispanic Black and Hispanic women, who represented more than 75% of the sample. This contradicts prior studies and indicates that although we trained the model with the majority of individuals from unprivileged groups with similar electronic medical record quality, the model was still biased.

Multiple reasons might explain our observed results. For instance, due to cultural differences, social stigma, or medical mistrust, non-Hispanic Black women might underreport their symptoms more often than non-Hispanic White women. Studies indicate that urban low-income women of color report lower levels of depressive symptoms than non-Hispanic White women using self-report questionnaires despite having the same diagnosis when assessed by a clinical provider (98). Thus, a lesser number of non-Hispanic Black women with prenatal depression might be referred to mental health providers. Our results agree with this. While our model identified non-Hispanic Black women with elevated depressive symptoms with a sensitivity of 80%, the positive predictive value was low (PPV=37%, **Figure S3d, g**). In other words, the model predicted a large proportion of non-Hispanic Black women as cases instead of control, indicating that a larger proportion of non-Hispanic Black women may have endorsed higher levels of depressive symptoms. However, our results are based on self-report screening tools and should be confirmed using clinical diagnosis. Another plausible reason for the lower predictive performance of our EMR-based ML model for women of color might be due to the different use of medical services. Despite all the EMR employed in this study being collected in the same clinic, non-Hispanic Black women, who were mostly in federal aid insurance (**Figure S2b**), had their first obstetric visit later in their pregnancy (59% after the first trimester vs. 51% within the first trimester) compared with non-Hispanic White women (6% after the first trimester vs. 11% within the first trimester) (p-value < 0.001, **Figure S2a**). Another possibility is that EMRs might not contain all the necessary information to predict prenatal depression, as some other relevant factors, such as chronic stress, are not measured or recorded in them. Certain neighborhood characteristics, such as violence, living in poverty areas, or air pollution, are known to increase chronic stress thus chronic inflammation (99) and inflammation is one of the hallmarks of perinatal depression (100). Thus, chronic stress and chronic inflammation might mediate the negative effects of structural inequalities in depression during pregnancy. Therefore, future research should include neighborhood factors that could boost model prediction performance in women who are exposed to higher contextual risks, e.g., racial minority and low-income women living in highly segregated urban poverty areas (101).

Our study has several strengths, including 1) the exploration of the predictive capabilities of ML models using EMR to identify depression early in pregnancy in low-income women of color, as most of the current studies aimed to predict postpartum depression in middle-class non-Hispanic White women; 2) the use of SHAP analysis to make model results interpretable and thus actionable; and 3) assessing model biases. Future studies can benefit from larger sample sizes in more diverse populations at high risk of PND (e.g., immigrants, homeless); using clinical diagnoses of prenatal depression, such as ICD-9/10 codes, instead of self-report depression questionnaires; the addition of community-level information; and the explorations of algorithms that could aid to remove model biases.

## Conclusions

Interpretable machine learning models based on available electronic medical records can aid in identifying women at high risk of PND in their early pregnancy so that clinicians can intervene early enough to prevent the negative consequences of PND for both the mother and the child. However, new tools and approaches are necessary to increase the prediction performance of EMR-based machine learning models to reduce model biases so that the risk of PND can be equitably predicted for all pregnant individuals.

## Acknowledgments

This work is funded through a K12 BIRCWH Award (NICHD 101373-04) BPB and supported by the National Center for Advancing Translational Sciences (NCATS), National Institutes of Health, through Grant Award Number UL1TR002003. We would like to thank Dr. Subhash Kumar Kolar Rajanna for his assistance in extracting the electronic medical records used in this manuscript. The content is solely the responsibility of the authors and does not necessarily represent the official views of the NIH.

## Data availability

Data is available to the research community upon approval of the University of Illinois Chicago Institutional Review Board (IRB # 2020-0553). Code is available at https://github.com/Bealab.

## Author contributions

BPB and YD conceptualized the idea; BPB, YD, and YH designed the methodology for model creation and validation; BPB, YH, and SA curated and pre-processed the data; YH performed the analysis and subsequent validation; BPB, YD, and YH interpreted the results and wrote the original draft; and all the authors critically reviewed and edited the manuscript.

