## Supplemental Figure for "Predicting prenatal depression and assessing model bias using machine learning models"

### Slide 1
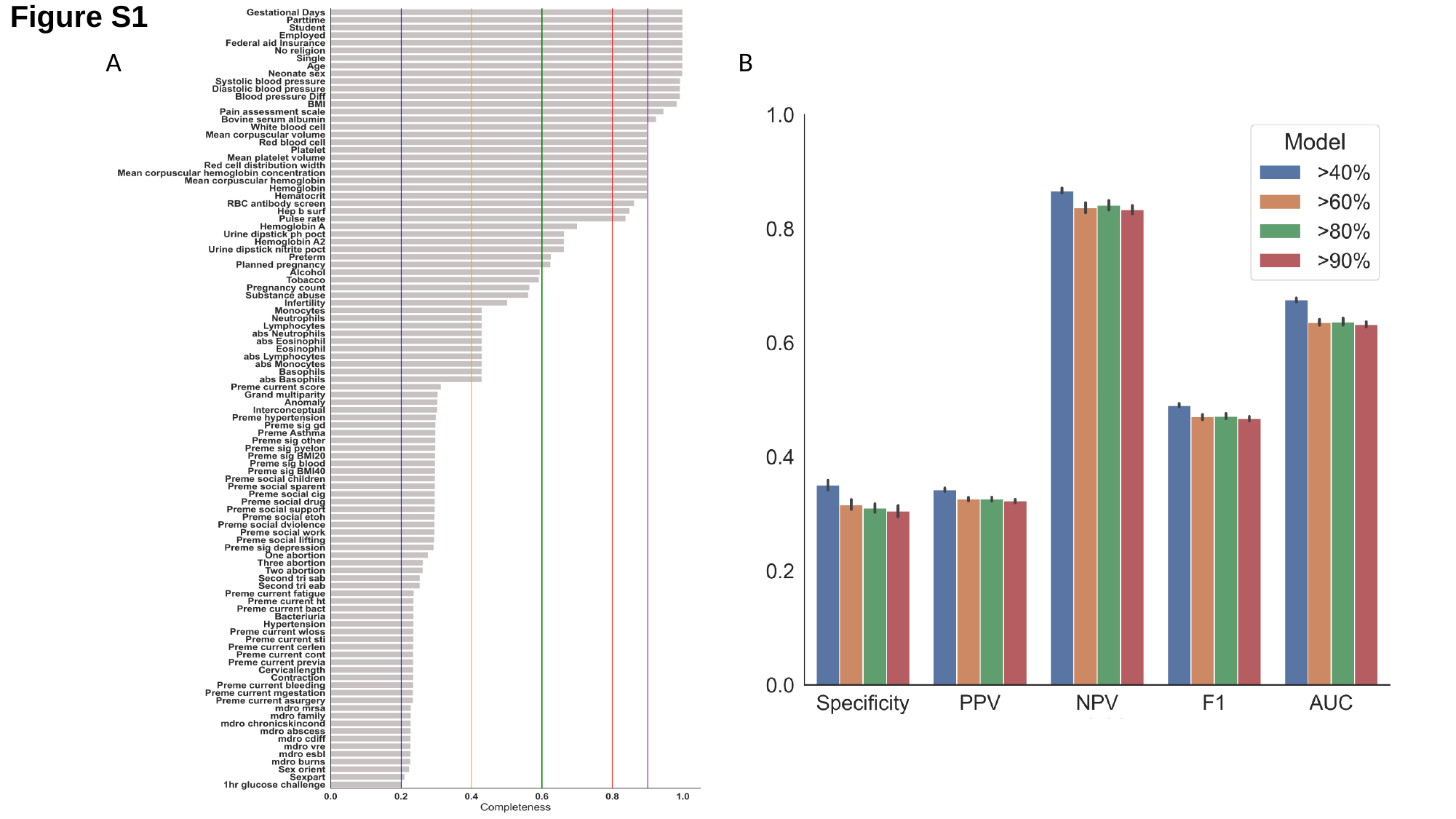

Figure S1
A
B

### Slide 2
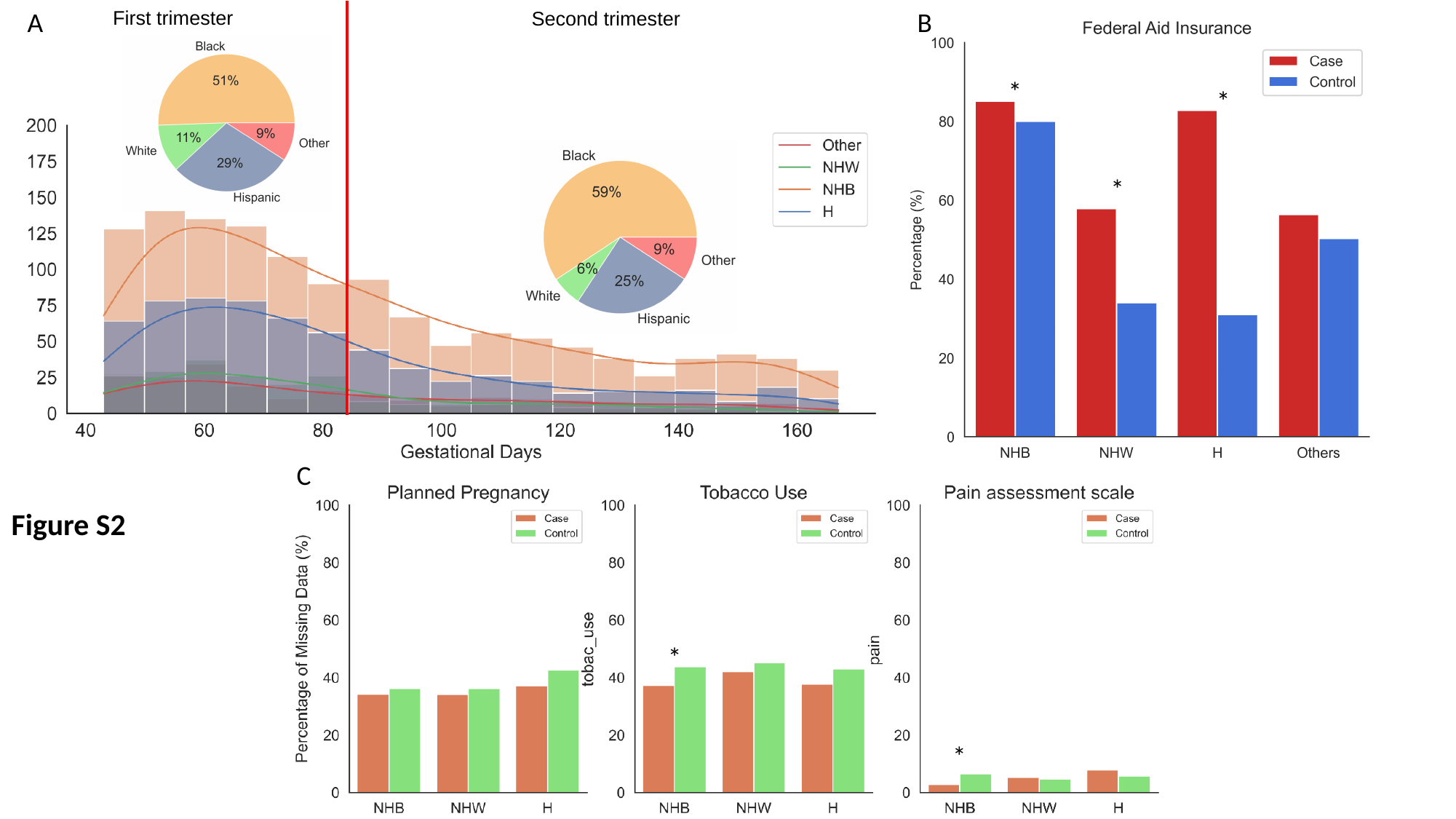

A
First trimester
B
Second trimester
*
*
*
C
Figure S2
*
*

### Slide 3
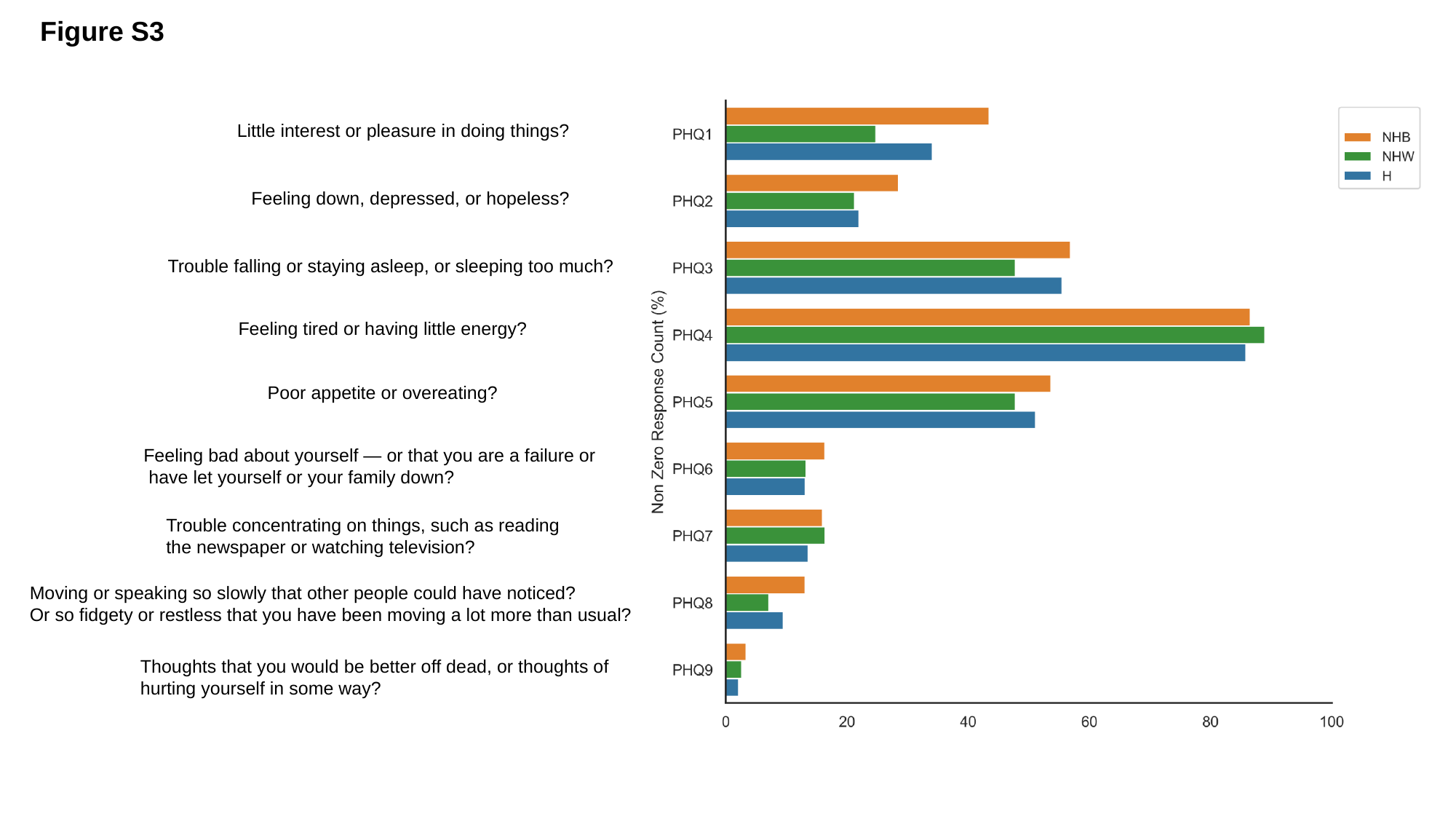

Figure S3
Little interest or pleasure in doing things?
Feeling down, depressed, or hopeless?
Trouble falling or staying asleep, or sleeping too much?
Feeling tired or having little energy?
Poor appetite or overeating?
Feeling bad about yourself — or that you are a failure or
 have let yourself or your family down?
Trouble concentrating on things, such as reading
the newspaper or watching television?
Moving or speaking so slowly that other people could have noticed?
Or so fidgety or restless that you have been moving a lot more than usual?
Thoughts that you would be better off dead, or thoughts of
hurting yourself in some way?

### Slide 4
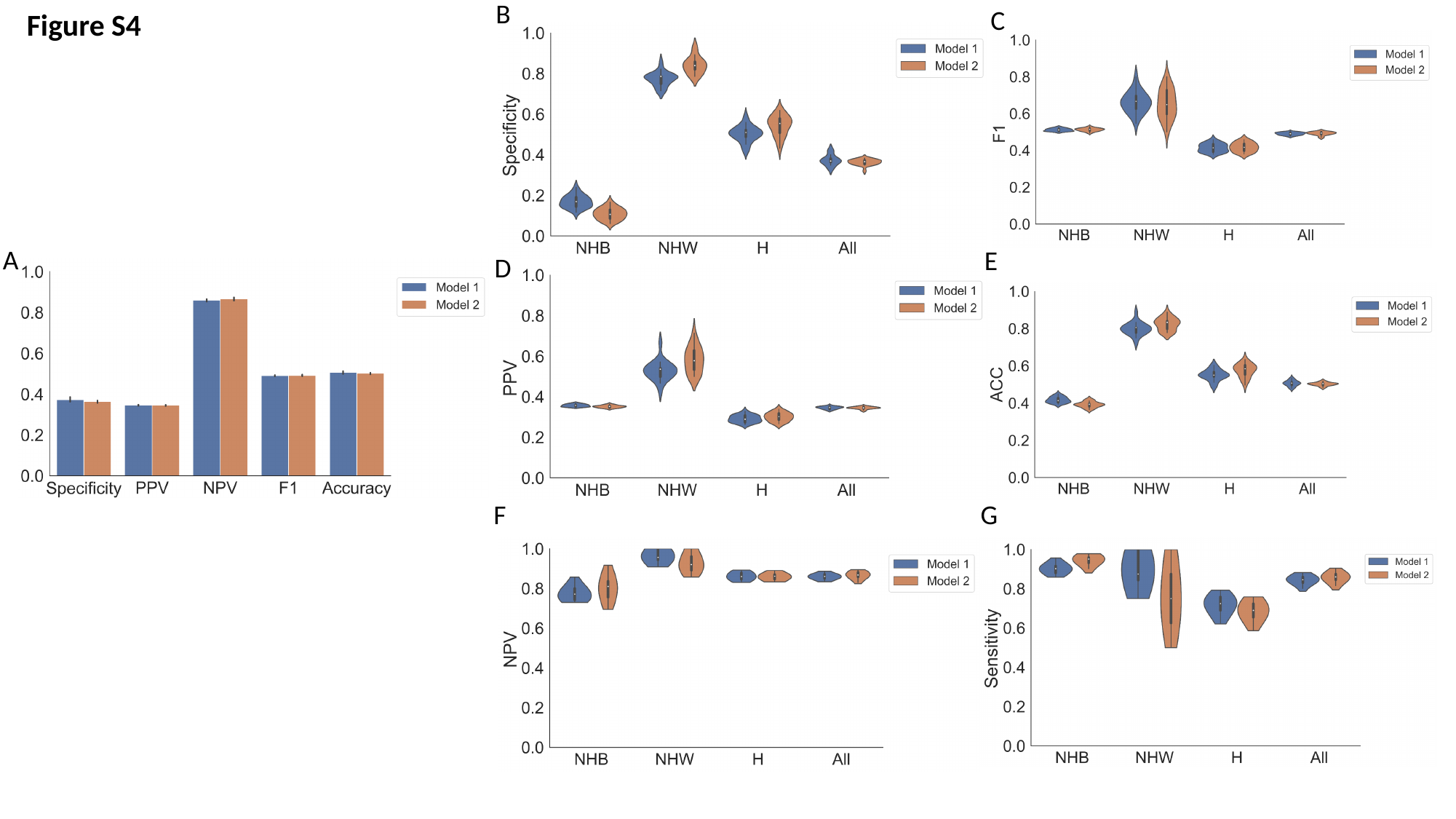

Figure S4
B
C
A
E
D
G
F

### Slide 5
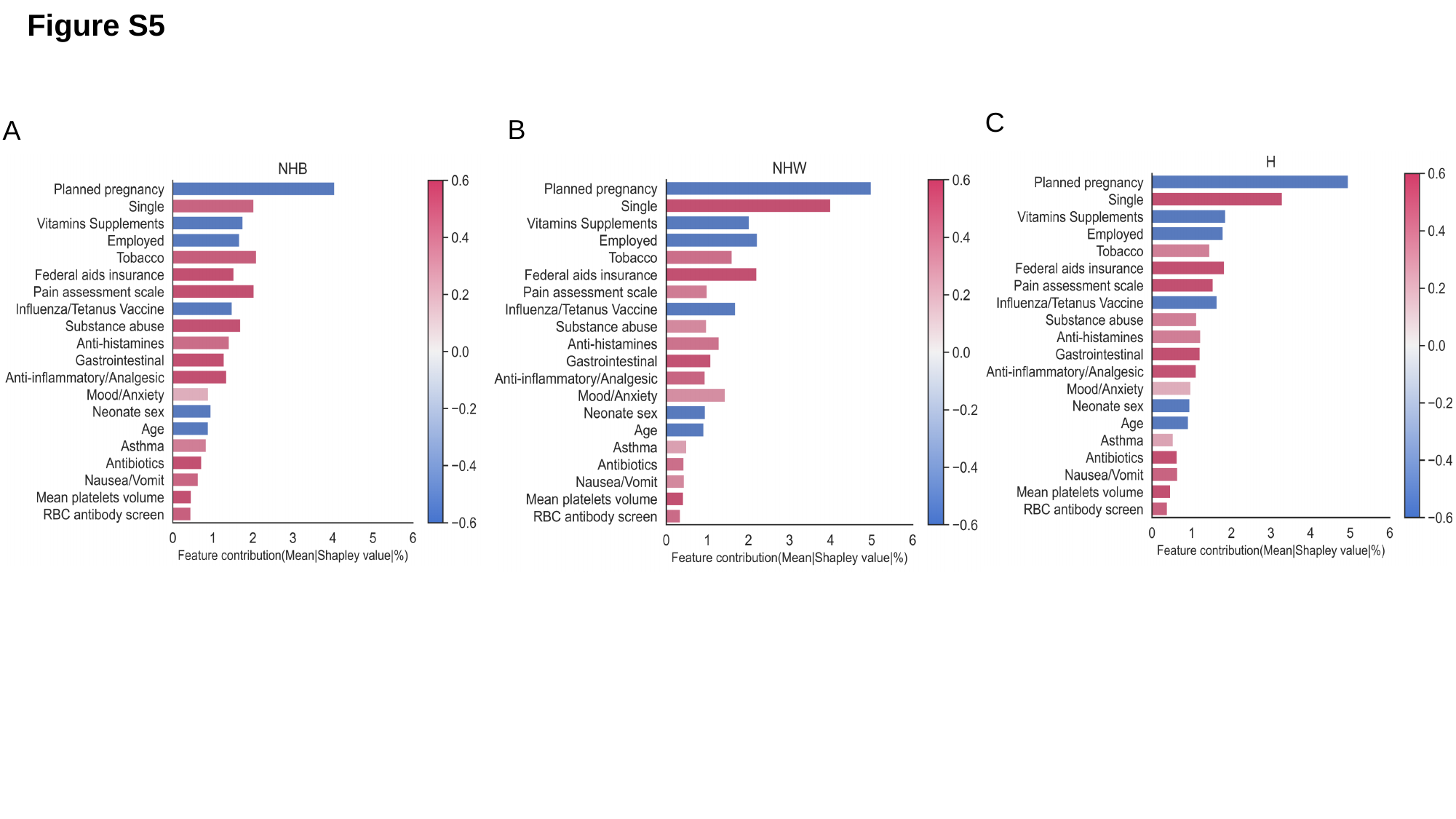

Figure S5
C
B
A

### Slide 6
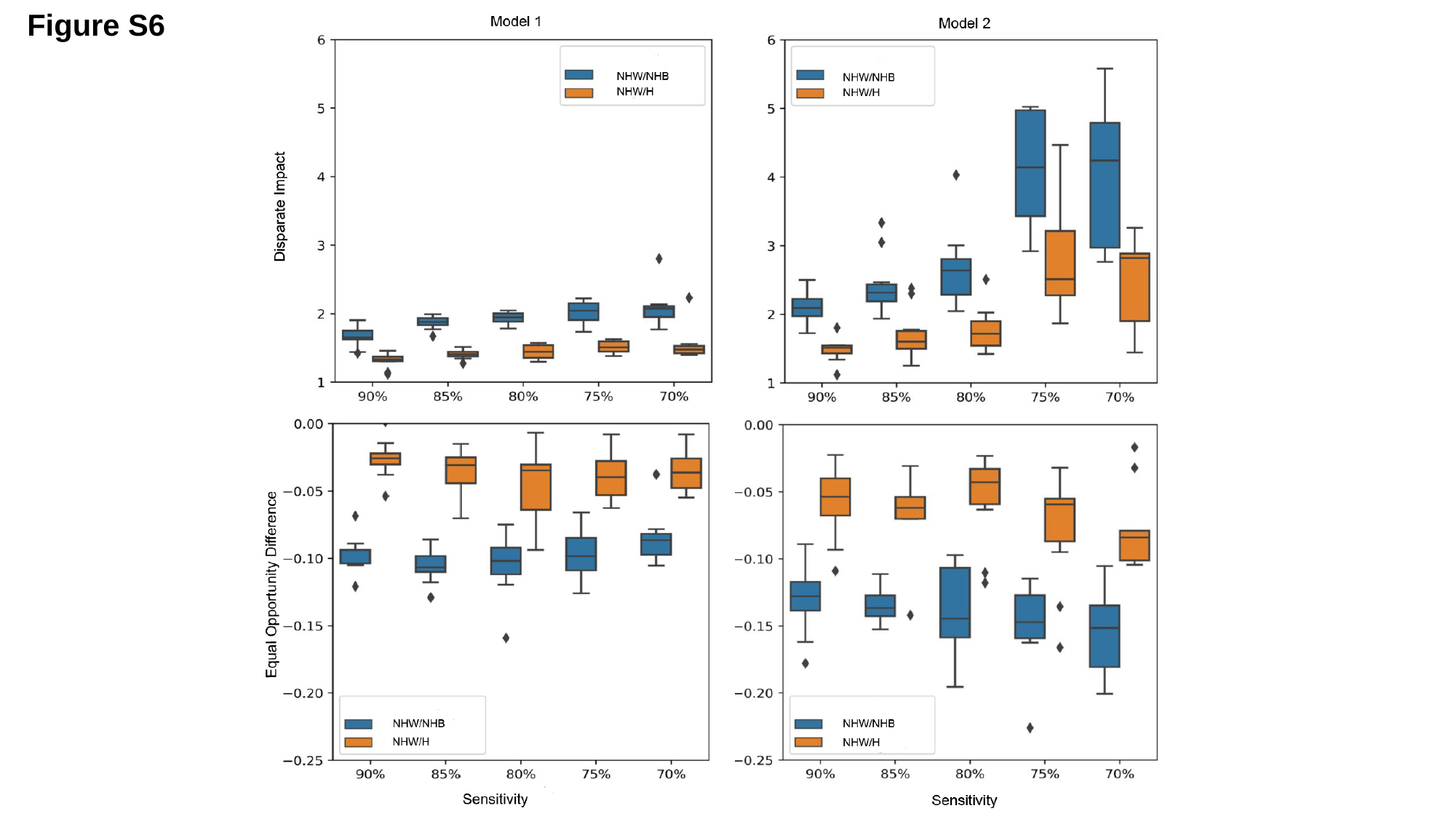

Figure S6
