## Supplements Caption for "Predicting prenatal depression and assessing model bias using machine learning models"

**SUPPLEMENT FIGURE LEGENDS**

**Supplement Table 1. Data dictionary.** The dictionary for all EMR features.

**Supplement Table 2. Medication.** The dictionary for all distinct medication.

**Supplement Table 3. Pair-wise comparison of social demographic features among races within positive or negative group.** Comparison of among races for each feature. Asterisk annotates significant differences (p adj < 0.05).

**Supplement Table 4. Pair-wise comparison of social demographic features within races.** The comparison within races for each feature. Asterisk annotates significant differences (p adj < 0.05).

**Supplement Table 5. Model performance.** Comparison of different machine learning methods.

**Supplement Figure 1. Sensitivity analysis of the imputation result. a,** completeness of data for each feature. **b,** summary statistics for model trained with different set of imputed data. Color represents the threshold of data completeness.

**Supplement Figure 2. Accessibility to care and the data missingness of top features. a,** distribution of gestational days across different race groups. The pie chart represents the distribution of race before and after 84 days of pregnancy. **b,** the percentage of individuals who has federal aid insurance across difference race with color represents case or control. **c,** the percentage of missing data in pain assessment scale, tobacco use and planned pregnancy across different race/ethnicity. NHB: non-Hispanic Black. NHW: non-Hispanic White. H: Hispanic or Latina.

**Supplement Figure 3. PHQ-9 response distribution.** The distribution of PHQ-9 response across different race groups. Each bar represents the percentage of score greater than 0. NHB: non-Hispanic Black. NHW: non-Hispanic White. H: Hispanic or Latina.

**Supplement Figure 4. Summary statistics for the model.** The orange and blue color indicates the model trained with or without race. **a,** The specificity, PPV, NPV, F1 and accuracy for all. **b,** Specificity in different race/ethnicity. **c,** F1 score in in different race/ethnicity. **d,** PPV in different race/ethnicity. **e,** Accuracy in different race/ethnicity. **f,** NPV in different race/ethnicity **g,** Sensitivity in different race/ethnicity. NHB: non-Hispanic Black. NHW: non-Hispanic White. H: Hispanic or Latina.

**Supplement Figure 5. Feature importance for difference race. a,** feature contribution for Non-Hispanic Black. **b,** feature contribution for Non-Hispanic White **c,** feature contribution for Hispanic.

**Supplement Figure 6. Sensitivity analysis on race disparity.** Disparate impact and equal opportunity difference of the model across different sensitivity threshold. NHB: non-Hispanic Black. NHW: non-Hispanic White. H: Hispanic or Latina.
